# A genome wide association study of frozen shoulder identifies a common variant of *WNT7B* and diabetes as causal risk factors

**DOI:** 10.1101/2020.11.13.20224360

**Authors:** Harry D Green, Alistair Jones, Jonathan P Evans, Andrew R Wood, Robin N Beaumont, Jessica Tyrrell, Timothy M Frayling, Chris Smith, Michael N Weedon

## Abstract

Frozen shoulder is a painful condition that often requires surgery and affects up to 5% of individuals aged 40-60 years. Little is known about the causes of the condition, but diabetes is a strong risk factor. To begin to understand the biological mechanisms involved, we aimed to identify genetic variants associated with frozen shoulder and to use Mendelian randomization to test the causal role of diabetes.

We performed a genome wide association study (GWAS) of frozen shoulder in the UK Biobank using data from 2064 cases identified from ICD-10 codes. We used data from FinnGen for replication. We used one-sample and two-sample Mendelian randomization approaches to test for a causal association of diabetes with frozen shoulder.

We identified a single genome-wide significant locus (lead SNP rs62228062; OR=1.34 [1.28-1.41], p=2×10^−16^) that contained *WNT7B*. A recent transcriptome study identified *WNT7B* as amongst the most enriched transcripts in anterior capsule tissue in patients undergoing arthroscopic capsulotomy surgery for frozen shoulder suggesting *WNT7B* as a potential causal gene at the locus. The lead SNP was also strongly associated with Dupuytren’s contracture (OR=2.61 [2.50, 2.72], p<1×10^−100^). The Mendelian randomization results provided evidence that type 1 diabetes is a causal risk factor for frozen shoulder (OR=1.04 [1.02-1.07], p=6×10^−5^). There was no evidence that obesity was causally associated with frozen shoulder, suggesting that diabetes influences risk of the condition through glycemic rather than mechanical effects.

We have identified the first genetic variant associated with frozen shoulder. *WNT7B* is a potential causal gene at the locus. Diabetes is a likely causal risk factor. Our results provide evidence of biological mechanisms involved in this common painful condition.

## Background

Frozen shoulder, also known as adhesive capsulitis, affects 2-5% of the population at some point in their lives [1, 2]. It is characterised by initial shoulder pain followed by a gradual reduction in range of movement or “freezing”. Pain subsides but joint stiffness can continue for years, causing significant disability [3]. The onset of frozen shoulder is usually between 40 and 60 years [4].

Little is known about the causes of frozen shoulder. Genome-wide association studies can provide new insights into underlying biological mechanism and possible drug targets. For example, a recent GWAS of osteoarthritis has been used to identify new therapeutic targets for the condition [5]. To date, there have been no published genome wide association studies (GWAS) of frozen shoulder.

Diabetes is the strongest known risk factor for diabetes. Individuals with diabetes have a greatly increased lifetime risk, with a hazard ratio of 1.33 [6]. Whether diabetes causes frozen shoulder is unclear because the association may reflect residual confounding by other risk factors such as age, obesity [7] and Dupuytren’s disease [8].

Mendelian randomization is a statistical method that can be used to infer causal relationships between an exposure and an outcome by using genetic variants associated with the exposure [9]. The exposure associated variants can be used as an unconfounded proxy for the exposure, as their inheritance is random at conception. This method is now extensively used to infer causal associations and proof of principle examples include evidence that increased BMI causes diabetes [10] and increased LDL cholesterol causes coronary artery disease [11].

Using the UK Biobank, a deeply phenotyped longitudinal study of 500,000 individuals from the UK, we performed a genome-wide association study (GWAS) to identify genetic variants associated with frozen shoulder. We used publicly available summary statistics from 135,658 individuals from the FinnGen study for replication [12]. We then used Mendelian randomization to test for causal associations of diabetes and obesity with frozen shoulder.

## Methods

### Frozen shoulder cases in the UK Biobank

The primary GWAS was performed on cases of frozen shoulder identified from the ICD-10 code M750 in the UK Biobank. For validation and sensitivity analyses in the UK Biobank primary care data we used read codes (N210, XE1FL and XE1Hm) and classified as cases any individual with at least one entry of these codes. Controls were defined as individuals without a record of one of these codes.

### Other phenotypes

ICD-10 codes were used for related disorders (Dupuytren’s disease (M720), any fibroblastic disease (M72), rotator cuff (M751), and calcific tendinitis of shoulder (M753)). Type 1 diabetes was classified favouring specificity over sensitivity, defined by diagnosis aged ≤ 20, on insulin within 1 year of diagnosis and at time of recruitment, not using oral antihyperglycemic agents, and not self-reporting type 2 diabetes, as described in [13]. Type 2 diabetes was defined by participants answering yes to the question ‘Has a doctor ever told you that you have diabetes?’ excluding those who reported using insulin within one year of diagnosis, were diagnosed under the age of 35, or were diagnosed within the past year [14]. We used UK Biobank variable 21001 for BMI measurement. HbA1c in mmol/mol is defined using variable n_30750_0_0 in UK Biobank. Diabetic eye disease was defined using self-report code 1276.

### Genome-wide Association Study

We performed a case-control genome-wide association study using BOLT-LMM [15], which applies a linear mixed model to test for an association between each SNP and the outcome trait, including population structure as a part of the model. Age, sex, study centre and genotyping chip were also included as covariates in the genome-wide association study. We performed a GWAS on 2,064 individuals in the UK Biobank classified as of European ancestry against 448,961 controls. Briefly, principal component analysis was performed using individuals from the 1000 Genomes Project prior to projection of UK Biobank individuals into the principal component space. K-means clustering was subsequently applied to classify individuals as European, with centres initiated to the mean principal component values of each 1000 Genomes sub-population. The first 4 principal components were used in this analysis. We used LocusZoom to plot the resulting significant locus [16] and GTEx to explore the possibility of functional variants [17].

### Replication analysis

We used summary statistics from freeze 3 of the FinnGen publicly available resource [12] to replicate the discovery GWAS findings. Cases of frozen shoulder were also defined using the ICD-10 code M750.

### Observational Analyses

We tested for associations with demographic and clinical features using logistic regression models, firstly using a univariable logistic regression model on each variable of interest and secondly using all variables in a multivariable logistic regression model to identify independent associations. We analysed Type 1 and Type 2 diabetes separately.

### Mendelian randomization

We applied two different methods. The one sample Mendelian randomization (MR) results were performed in two stages: first, the association between each exposure and a genetic risk score was used to derive a genetically predicted exposure value, and second, these predicted values were used in a logistic regression model of frozen shoulder, adjusting for age, sex, ancestral principal components, assessment centre and genotyping platform.

Second, we used two-sample methods, which involve regressing the effect sizes of variant– outcome associations against the effect sizes of the variant–risk factor associations for a set of SNPs associated with the exposure. We performed inverse variance weighted (IVW) instrumental variable analysis and two further methods that are more robust to potential violations of the standard MR assumptions (MR-Egger [18] and weighted-median MR [19]). We used effect sizes and SNPs for type 1 diabetes reported in [20] using the Wellcome Trust Case Control Consortium, and a type 2 diabetes genetic risk score using variants identified in [21], [22]. The T1D-GRS uses 2 SNPs, rs2187668 and rs7454108, to tag and categorize the high risk DR3/DR4 haplotypes [20], this requires access to individual level data which we did not have for the FinnGen study. We therefore performed two analyses: one where we used the 2 SNPs to impute DR3/DR4 status and another where we used the SNPs as standard risk SNPs.

## Results

### Diabetes is strongly associated with frozen shoulder in the UK Biobank

To identify associated risk factors for frozen shoulder we tested several measures of diabetes and its related traits, in univariable and, to assess their independent contributions, multivariable analyses. There were 2,064 cases of frozen shoulder in the UK Biobank based on ICD-10 codes and 7,913 based on GP records, of which 323 were common to both. Demographic and clinical associations with Frozen shoulder can be found in **Table 1**. Those with frozen shoulder were more likely female, had lower Townsend deprivation index, more likely obese (higher BMI and waist-hip-ratio) and more likely to have either type 1 or type 2 diabetes, with type 1 giving the strongest association. In a multivariable logistic regression model, only the type 1 and type 2 diabetes showed strong association.

**Table 1:**
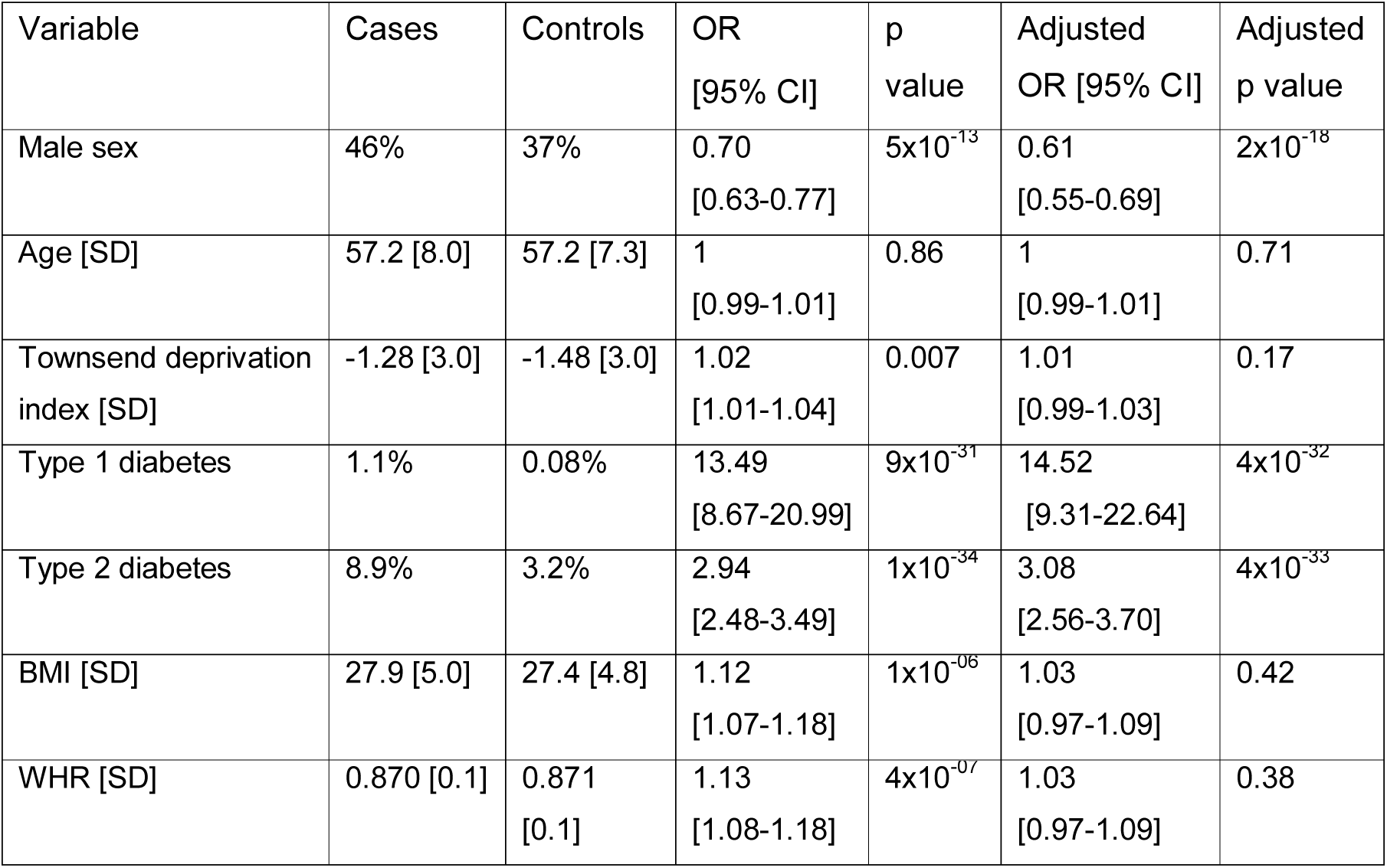
Demographic and clinical associations with frozen shoulder in the UK Biobank. The OR and p value columns were calculated using univariable logistic regression. The adjusted OR and adjusted p value columns were calculated using a multivariable logistic regression model. Adjusted results for diabetes types were calculated from a model excluding the other type.

Diabetes duration was associated with frozen shoulder (OR per additional year with diabetes 1.02 [1.01-1.04], p=0.005). HbA1c associated with frozen shoulder independently of type or duration of diabetes (OR=1.11 [95% CI: 1.04-1.20], p=0.002). Diabetic eye disease associated with frozen shoulder independently of type or duration of diabetes (OR = 1.84 [95% CI: 1.02-3.30], p=0.04). This suggests that individuals with longer duration and less well controlled diabetes have higher risk of diabetes.

### Variants in WNT7B are strongly associated with frozen shoulder

**Figure 1** presents the results of the frozen shoulder genome-wide association study. We identify a single genome-wide significant peak on chromosome 22. The G allele at the lead SNP, rs62228062, has a frequency of 31.4% in cases and 25.8% in controls (OR=1.34, [95% CI: 1.28, 1.41], p=2×10^−16^). **Figure 2** shows the Locuszoom [16] plot of the associated locus.

**Figure 1:**
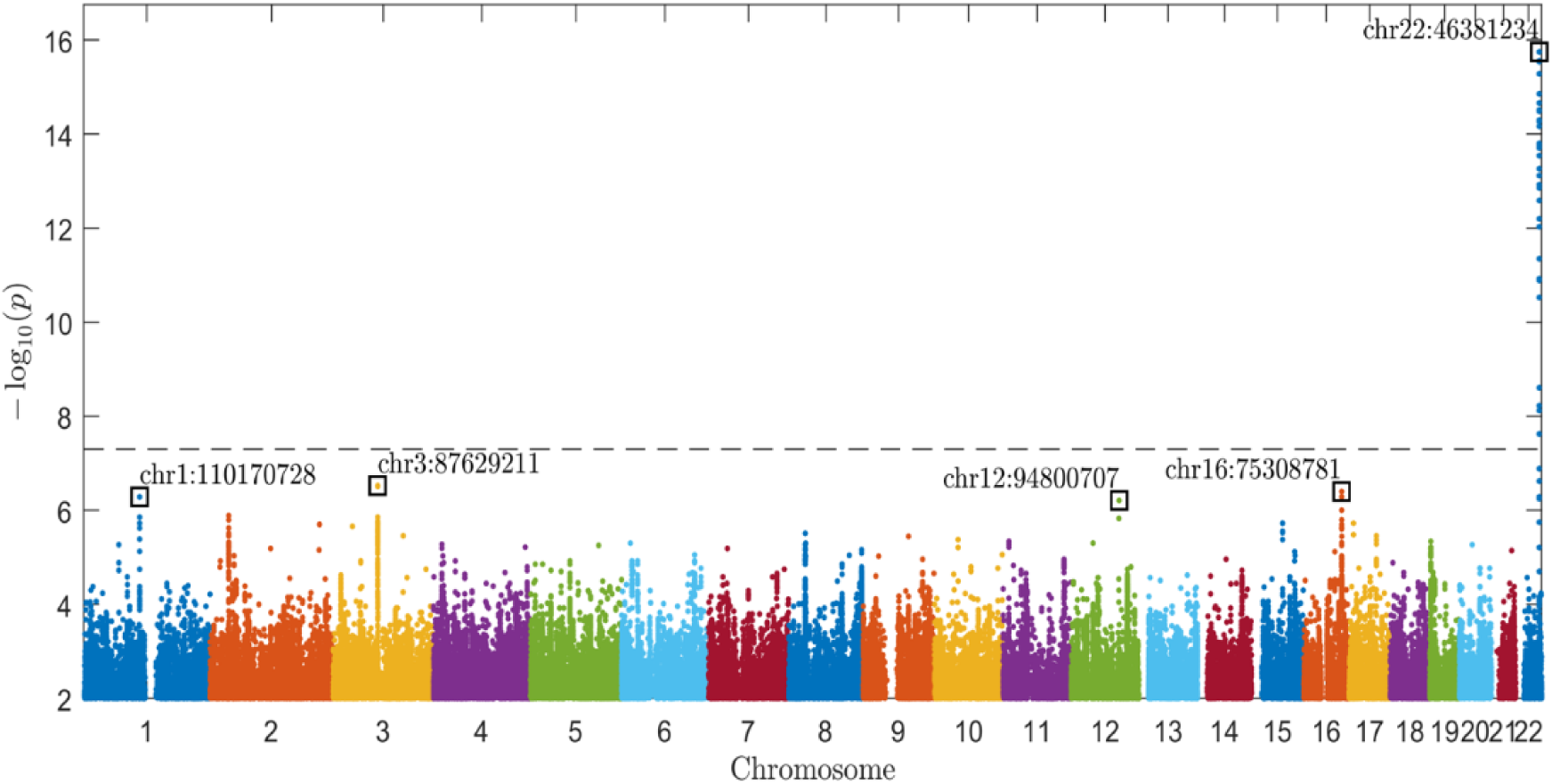
Manhattan plot of discovery GWAS for frozen shoulder in UK Biobank. The plot shows –log10(p) values for each single nucleotide polymorphism [SNP] in the HRC Imputation Panel with p < 0.01, computed using BOLT-LMM and plotted using an in-house MATLAB script which we have made publicly available on the MATLAB File Exchange [23]. The horizontal dashed line is the genome-wide significance threshold at p=5×10^−8^. Positions are based on the hg19 reference human genome.

**Figure 2:**
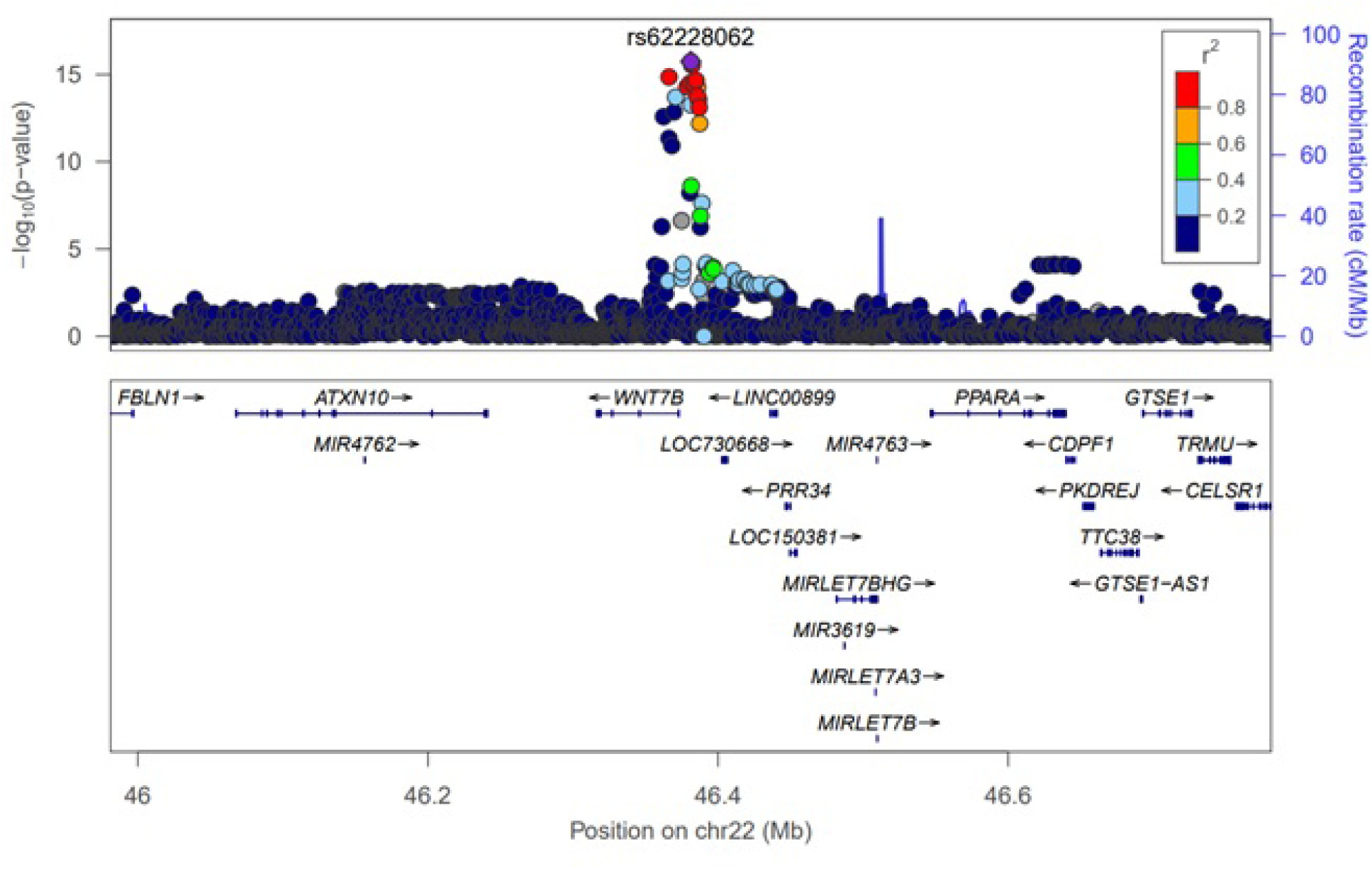
Locus Zoom plot. Showing the local LD structure and nearby genes to SNP rs62228062.

### Replication and sensitivity analyses of the WNT7B association signal

We sought replication of the association signal in the FinnGen study. FinnGen have recently released GWAS summary statistics for 1801 diseases and traits, including frozen shoulder [12]. The lead SNP from the UK Biobank, rs62228062, was associated with frozen shoulder in FinnGen with a similar effect size to UK Biobank (OR=1.29 [95% CI: 1.19, 1.40], p=2×10^−9^). The frequency of the G allele was 25.9% in cases compared to 21.9% in controls.

When using 7,913 cases identified only from primary care records in UK Biobank, rs62228062 associated with frozen shoulder, but with a substantially attenuated effect (OR=1.13 [95% CI: 1.09-1.17], p=2×10^−12^).

### WNT7B is the likely causal gene at the locus

There were no coding variants with an r^2^ > 0.8 with rs62228062. There are no gene expression associations in GTEX for rs62228062. However, a recent study found *WNT7B* was the second most differentially expressed transcript genome-wide with a log fold change of 7 (*p=*1×10^−16^) in anterior capsule tissue from 22 patients undergoing arthroscopic capsulotomy surgery for frozen shoulder compared to 26 controls [24].

### SNPs in the region have been associated with Dupuytren’s contracture and bone mineral density

A previous GWAS study of Dupuytren’s disease found an association in the same genomic region of *WNT7B* [25]. The lead SNP for Dupuytren’s contracture, rs7291412, was not associated with frozen shoulder in UK Biobank (OR=1.03 [95%CI: 0.96, 1.09], p=0.44) or FinnGen (OR=1.00 [95%: 0.93, 1.08], p=0.98). Our lead SNP, rs62228062, has previously been associated with heel bone mineral density [26] and we replicate that finding in UK Biobank with the G allele increasing bone mineral density (beta=0.003 [95% CI: 0.002, 0.003], p=2×10^−14^).

### The frozen shoulder SNP is also strongly associated with Dupuytren’s disease and other fibroblastic diseases

**Table 2** shows the association of rs62228062 with related disorders in UK Biobank. rs62228062 is associated strongly with Dupuytren’s disease, but not other related shoulder conditions. The association of rs62228062 with frozen shoulder did not change when we excluded individuals with Dupuytren’s disease from the cases (OR=1.31 [95%CI: 1.22-1.4], p=3×10^−15^)

**Table 2:**
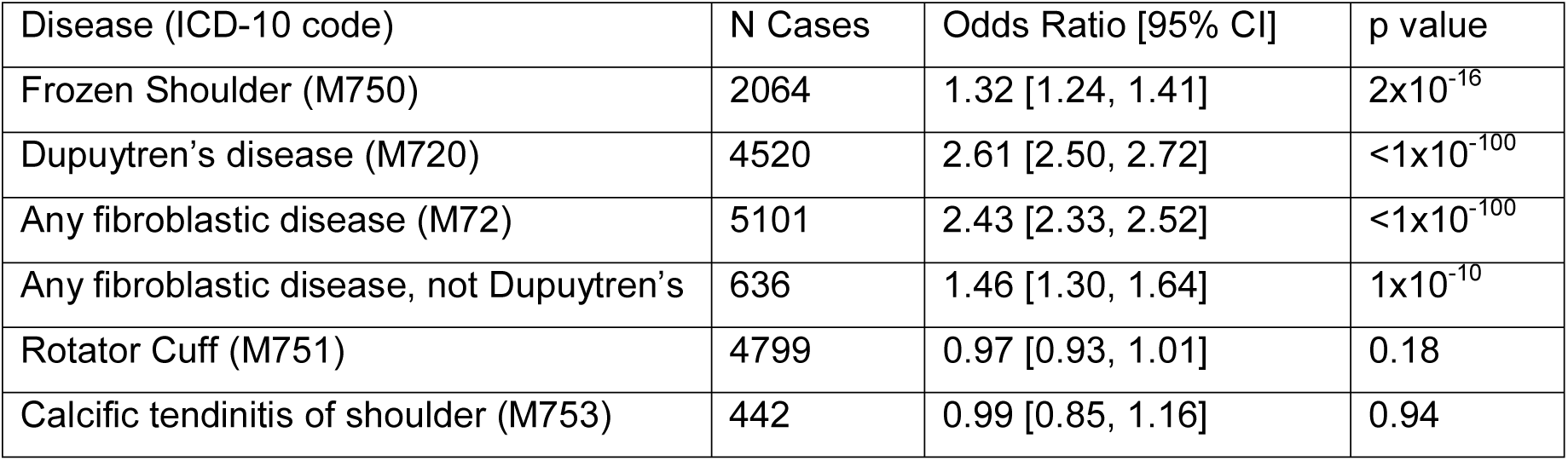
Association of lead SNP (rs62228062) against related disorders in UK Biobank.

### Mendelian randomization provides evidence that diabetes is causal to frozen shoulder

We used Mendelian randomization to explore whether the associations with Type 1 and Type 2 diabetes reported in **Table 2** were causal. Using 1-sample Mendelian randomization methods, genetic data provided evidence that type 1 diabetes causes frozen shoulder: OR 1.05 [95% CI: 1.01-1.09], p=0.01 using ICD-10 codes and OR 1.03 [95% CI: 1.02-1.05], p=4×10^−5^ including primary care records records. Evidence of a causal role of type 2 diabetes was weaker: OR 1.11 [95% CI: 0.99-1.24], p=0.08 using ICD-10 codes and OR 1.07 [95% CI: 1.02-1.13], p=0.008 including cases identified by UK Biobank primary care records.

The association with type 1 diabetes was replicated using more robust, two-sample MR methods IVW (p=6×10^−4^) and MR-Egger (p=2×10^−3^). There was no evidence from the two-sample methods that type 2 diabetes causes frozen shoulder (IVW: p=0.17, MR-Egger: p=0.45). The supplementary tables contain the full results from our Mendelian randomization analyses, including T1D-GRS results that exclude HLA variants which have greater potential for pleiotropy (**Supplementary Table 1** shows the two stage regression, **Supplementary Table 2** shows the IVW results and **Supplementary Table 3** shows the MR-Egger results). **Figure 3** shows a plot showing the lines of best fit for MR-Egger, IVW, Median IV and Penalised Median IV methods for type 1 and type 2 diabetes defined by ICD-10 codes, demonstrating a) for type 1 diabetes SNPs, a strong correlation between the betas for type 1 diabetes and frozen shoulder and b) no association for type 2 diabetes SNPs.

**Figure 3:**
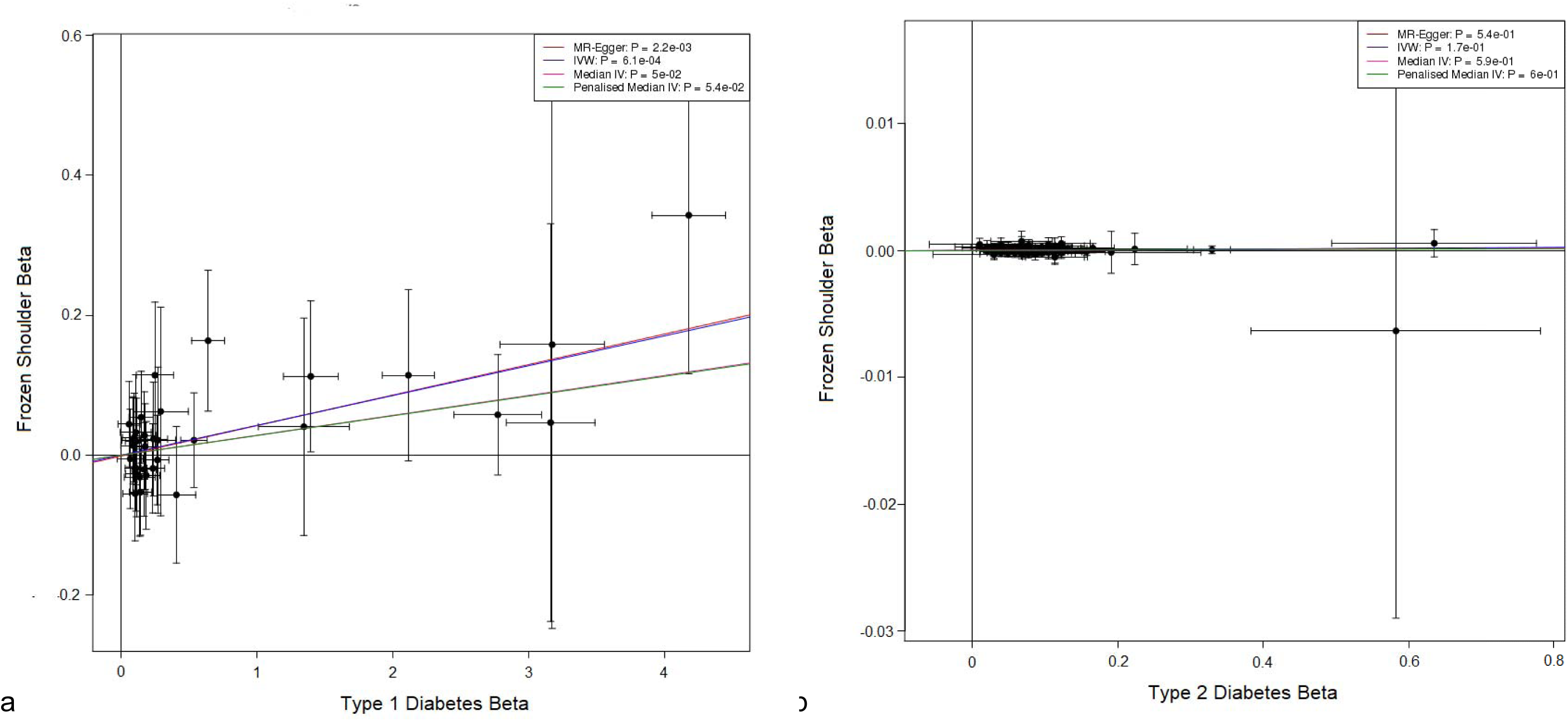
Two Sample Mendelian Randomization Results, showing for each SNP in the Type 1 Diabetes GRS in [20], the log-odds ratio for a) type 1 diabetes and b) type 2 diabetes on the x axis and the log-odds ratio for frozen shoulder (defined by ICD-10 code) on the y axis.

## Discussion

We have identified the first robust genetic variant to be associated with frozen shoulder. Frozen shoulder is a condition which affects up to 5% of the population around between the ages of 40 to 60, but the causes of the disease, and particularly the transient nature of the condition are unknown. The genome-wide association study and Mendelian randomization analyses we report here provide new insights into the underlying causes of the condition.

*WNT7B* is a strong candidate gene at the locus. As with most GWAS studies, further work is needed to identify the causal variant at the locus. GTEX analyses did not identify any strong eQTL’s with the lead SNP at the associated locus. However, a recent study performed RNA-seq on anterior capsule tissue from 22 patients undergoing arthroscopic capsulotomy surgery for adhesive capsulitis and compared to 26 undergoing arthroscopic stabilization surgery for a different condition [24]. *WNT7B* was the second most differentially expressed transcript genome-wide with a log fold change of 7 (*p=*1×10^−16^), although it was noted that the expression levels of *WNT7B* was relatively low. The WNT signalling pathway has been highlighted by previous GWAS of related fibroblastic diseases [27].

The lead variant also strongly associates with Dupuytren’s disease. Dupuytren’s disease is a common condition which is characterised by a hand deformity where fingers cannot be elongated fully. A previous GWAS of Dupuytren’s identified 26 associated loci with the condition [25]. One of the more strongly associated loci also contained the *WNT7B* locus. However, the lead SNP at that locus has not been associated with frozen shoulder and does not associate with frozen shoulder in the UK Biobank or FinnGen. The odds ratio for the lead SNP in our study has a significantly stronger odds ratio for Dupuytren’s than the strongest association signal from the Dupuytren’s GWAS. The explanation may be that older versions of SNP chips or imputation panels were used in the previous study which did not capture the lead SNP from our current study of frozen shoulder. It is also unclear why a variant that causes a chronic condition of Dupuytren’s disease would cause the late onset and transient condition of frozen shoulder.

Our analyses demonstrate that diabetes is a causal risk factor for frozen shoulder. Diabetes is known to be a strong observational risk factor for frozen shoulder, but the causal nature of the association was unclear. Our Mendelian randomization analyses in UK Biobank provide evidence that Type 1 diabetes is causal for the condition. The weaker association with Type 2 diabetes is likely due to differences in duration of diabetes as Type 1 diabetes is generally diagnosed earlier (half before the age of 30 years) whereas the diagnosis of Type 2 diabetes generally occurs later in life (after 50 years). It is likely all individuals with diabetes would be an increased risk of frozen shoulder, with those with longer duration (like those individuals with Type 1 diabetes) and worse glycemic control at increased risk.

There are limitations to our study. The UK Biobank ICD-10 data is reliant on patients having a coded diagnosis at hospital, which is reliant on accuracy of hospital coding, and also reliant on the condition being severe enough for the patient to go to hospital, which may result in less serious cases being classified as controls. However, frozen shoulder can be difficult to accurately diagnose in primary care. Consistent with this the effect size for frozen shoulder was smaller when cases were based on presence in primary care records rather than inpatient ICD-10 codes. This study was performed using only white Europeans, and further study is needed to determine if results replicate for other ethnic groups. The UK Biobank only includes patients between the ages of 40 and 69 at recruitment, although this covers the most common incidence age range for frozen shoulder.

In conclusion, we have identified the first genetic variant associated with frozen shoulder. *WNT7B* is the likely causal gene at the locus. We demonstrate that diabetes is a causal risk factor for frozen shoulder.

## Data Availability

This research has been conducted using the UK Biobank Resource (Application number: 68492).

## Acknowledgements

This research has been conducted using the UK Biobank Resource (Application number: 68492). The Genotype-Tissue Expression (GTEx) Project was supported by the <u>Common Fund</u> of the Office of the Director of the National Institutes of Health, and by NCI, NHGRI, NHLBI, NIDA, NIMH, and NINDS. “We want to acknowledge the participants and investigators of the FinnGen study”. HG was funded by an “Expanding excellence in England” award from Research England”. TMF has received funding from the Medical Research Council, MR/T002239/1 and the Innovative Medicines Initiative 2 Joint Undertaking under grant agreement No 875534. This Joint Undertaking support from the European Union’s Horizon 2020 research and innovation programme and EFPIA and T1D Exchange, JDRF, and Obesity Action Coalition.

## References

[1] I. Morén-Hybbinette, U. Moritz, and B. Scherstén, “The clinical picture of the painful diabetic shoulder--natural history, social consequences and analysis of concomitant hand syndrome.,” Acta Med. Scand., vol. 221, no. 1, pp. 73–82, 1987, doi: 10.1111/j.0954-6820.1987.tb01247.x.

[2] N. H. Zreik, R. A. Malik, and C. P. Charalambous, “Adhesive capsulitis of the shoulder and diabetes: a meta-analysis of prevalence.,” Muscles. Ligaments Tendons J., vol. 6, no. 1, pp. 26–34, 2016, doi: 10.11138/mltj/2016.6.1.026.

[3] C. Hand, K. Clipsham, J. L. Rees, and A. J. Carr, “Long-term outcome of frozen shoulder.,” J. shoulder Elb. Surg., vol. 17, no. 2, pp. 231–236, 2008, doi: 10.1016/j.jse.2007.05.009.

[4] H. S. Uppal, J. P. Evans, and C. Smith, “Frozen shoulder: A systematic review of therapeutic options,” World J. Orthop., vol. 6, no. 2, pp. 263–268, 2015, doi: 10.5312/wjo.v6.i2.263.

[5] I. Tachmazidou et al., “Identification of new therapeutic targets for osteoarthritis through genome-wide analyses of UK Biobank data.,” Nat. Genet., vol. 51, no. 2, pp. 230–236, Feb. 2019, doi: 10.1038/s41588-018-0327-1.

[6] Y.-P. Huang et al., “Association of diabetes mellitus with the risk of developing adhesive capsulitis of the shoulder: a longitudinal population-based followup study.,” Arthritis Care Res. (Hoboken)., vol. 65, no. 7, pp. 1197–1202, Jul. 2013, doi: 10.1002/acr.21938.

[7] K. Kingston, E. J. Curry, J. W. Galvin, and X. Li, “Shoulder adhesive capsulitis: epidemiology and predictors of surgery.,” J. shoulder Elb. Surg., vol. 27, no. 8, pp. 1437–1443, Aug. 2018, doi: 10.1016/j.jse.2018.04.004.

[8] S. P. Smith, V. S. Devaraj, and T. D. Bunker, “The association between frozen shoulder and Dupuytren’s disease.,” J. shoulder Elb. Surg., vol. 10, no. 2, pp. 149–151, 2001, doi: 10.1067/mse.2001.112883.

[9] N. M. Davies, M. V Holmes, and G. Davey Smith, “Reading Mendelian randomisation studies: a guide, glossary, and checklist for clinicians,” BMJ, vol. 362, p. k601, Jul. 2018, doi: 10.1136/bmj.k601.

[10] L. J. Corbin et al., “BMI as a modifiable risk factor for type 2 diabetes: Refining and understanding causal estimates using mendelian randomization,” Diabetes, vol. 65, no. 10, pp. 3002–3007, 2016, doi: 10.2337/db16-0418.

[11] M. V Holmes et al., “Mendelian randomization of blood lipids for coronary heart disease.,” Eur. Heart J., vol. 36, no. 9, pp. 539–550, Mar. 2015, doi: 10.1093/eurheartj/eht571.

[12] FinnGen, “FinnGen Documentation of R3 release,” 2020. [Online]. Available: https://finngen.gitbook.io/documentation/. [Accessed: 26-Oct-2020].

[13] S. A. Sharp et al., “Development and Standardization of an Improved Type 1 Diabetes Genetic Risk Score for Use in Newborn Screening and Incident Diagnosis,” vol. 42, no. February, pp. 200–207, 2019, doi: 10.2337/dc18-1785.

[14] J. S. Tyrrell, H. Yaghootkar, R. M. Freathy, A. T. Hattersley, and T. M. Frayling, “Parental diabetes and birthweight in 236 030 individuals in the UK Biobank Study,” pp. 1714–1723, 2013, doi: 10.1093/ije/dyt220.

[15] P.-R. Loh, G. Kichaev, S. Gazal, A. P. Schoech, and A. L. Price, “Mixed-model association for biobank-scale datasets.,” Nature genetics, vol. 50, no. 7. pp. 906–908, Jul-2018, doi: 10.1038/s41588-018-0144-6.

[16] R. J. Pruim et al., “LocusZoomD: regional visualization of genome-wide association scan results,” vol. 26, no. 18, pp. 2336–2337, 2010, doi: 10.1093/bioinformatics/btq419.

[17] “The GTEx Consortium atlas of genetic regulatory effects across human tissues.,” Science, vol. 369, no. 6509, pp. 1318–1330, Sep. 2020, doi: 10.1126/science.aaz1776.

[18] J. Bowden, G. D. Smith, and S. Burgess, “Mendelian Randomization Methodology Mendelian randomization with invalid instrumentsD: effect estimation and bias detection through Egger regression,” no. June, pp. 512–525, 2015, doi: 10.1093/ije/dyv080.

[19] J. Bowden, G. Davey Smith, P. C. Haycock, and S. Burgess, “Consistent Estimation in Mendelian Randomization with Some Invalid Instruments Using a Weighted Median Estimator,” Genet. Epidemiol., vol. 40, no. 4, pp. 304–314, May 2016, doi: 10.1002/gepi.21965.

[20] R. A. Oram et al., “A type 1 diabetes genetic risk score can aid discrimination between type 1 and type 2 diabetes in young adults,” Diabetes Care, vol. 39, no. 3, pp. 337–344, 2016, doi: 10.2337/dc15-1111.

[21] A. Mahajan et al., “Genome-wide trans-ancestry meta-analysis provides insight into the genetic architecture of type 2 diabetes susceptibility.,” Nat. Genet., vol. 46, no. 3, pp. 234–244, Mar. 2014, doi: 10.1038/ng.2897.

[22] A. P. Morris et al., “Large-scale association analysis provides insights into the genetic architecture and pathophysiology of type 2 diabetes.,” Nat. Genet., vol. 44, no. 9, pp. 981–990, Sep. 2012, doi: 10.1038/ng.2383.

[23] H. D. Green, “Manhattan Plots for visualisation of GWAS results,” A function for plotting a Manhattan Plot in MATLAB directly from a text file of GWAS statistics from PLINK, BOLT-LMM or SAIGE. GB, 01-Mar-2019.

[24] N. Kamal et al., “Transcriptomic analysis of adhesive capsulitis of the shoulder.,” J. Orthop. Res. Off. Publ. Orthop. Res. Soc., vol. 38, no. 10, pp. 2280–2289, Oct. 2020, doi: 10.1002/jor.24686.

[25] M. Ng et al., “A Genome-wide Association Study of Dupuytren Disease Reveals 17 Additional Variants Implicated in Fibrosis,” Am. J. Hum. Genet., vol. 101, no. 3, pp. 417–427, 2017, doi: 10.1016/j.ajhg.2017.08.006.

[26] S. K. Kim, “Identification of 613 new loci associated with heel bone mineral density and a polygenic risk score for bone mineral density, osteoporosis and fracture,” PloS One, vol. 13, no. 7, p. e0200785, 2018, doi: 10.1371/journal.pone.0200785.

[27] E.-J. P. M. ten Dam, M. M. van Beuge, R. A. Bank, and P. M. N. Werker, “Further evidence of the involvement of the Wnt signaling pathway in Dupuytren’s disease,” J. Cell Commun. Signal., vol. 10, no. 1, pp. 33–40, 2016, doi: 10.1007/s12079-015-0312-8.

