## Supplementary Table for "A genome wide association study of frozen shoulder identifies a common variant of *WNT7B* and diabetes as causal risk factors"

### Supplementary Tables

#### Supplementary Table 1 – One Sample Mendelian Randomisation Results

| Exposure | Outcome | OR | P value |
|---|---|---|---|
| T1D-GRS | FS_ICD-10 | 1.04 (1.00-1.08) | 0.03 |
| T1D-GRS | FS_GP | 1.03 (1.02-1.05) | 4e-5 |
| T2D GRS | FS_ICD-10 | 1.11 (0.99-1.24) | 0.08 |
| T2D GRS | FS_GP | 1.07 (1.02-1.13) | 0.008 |

#### Supplementary Table 2 – IVW Mendelian Randomisation Results

| Exposure | Outcome | OR | P value |
|---|---|---|---|
| T1D | UKBB ICD-10 | 1.04 (1.02-1.07) | 6e-5 |
| T1D no DR3/DR4 haplotyping | UKBB ICD-10 | 1.04 (1.01-1.07) | 0.003 |
| T1D no DR3/DR4 haplotyping | UKBB GP | 1.03 (1.02-1.04) | 1e-4 |
| T1D no DR3/DR4 haplotyping | FinnGen | 1.07 (1.04-1.11) | 8e-5 |
| T1D no DR3/DR4 haplotyping | Meta Analysis | 1.05 (1.03-1.08) | 0.000 |
| T1D no HLA | UKBB ICD-10 | 1.06 (0.98-1.14) | 0.15 |
| T1D no HLA | FinnGen | 1.10 (1.02-1.18) | 0.02 |
| T1D no HLA | Meta Analysis | 1.08 (1.03-1.14) | 0.004 |
| T2D | UKBB ICD-10 | 1.00 (1.00-1.00) | 0.17 |
| T2D | UKBB GP | 1.03 (1.00-1.07) | 0.05 |
| T2D | FinnGen | 0.99 (0.90-1.09) | 0.80 |
| T2D | Meta Analysis | 1.00 (1.00-1.00) | 0.83 |

Supplementary Table 3 – MR-Egger Results

| Exposure | Outcome | OR | P value | p int |
|---|---|---|---|---|
| T1D | UKBB ICD-10 | 1.04 (1.02-1.07) | 0.002 | 0.88 |
| T1D no DR3/DR4 haplotyping | UKBB ICD-10 | 1.04 (1.01-1.07) | 0.02 | 0.96 |
| T1D no DR3/DR4 haplotyping | UKBB GP | 1.03 (1.02-1.05) | 1e-4 | 0.25 |
| T1D no DR3/DR4 haplotyping | FinnGen | 1.07 (1.03-1.11) | 0.002 | 0.47 |
| T1D no DR3/DR4 haplotyping | Meta Analysis | 1.05 (1.03-1.08) | 0.000 | |
| T1D no HLA | UKBB ICD-10 | 1.10 (0.97-1.26) | 0.16 | 0.46 |
| T1D no HLA | FinnGen | 1.09 (0.96-1.24) | 0.20 | 0.87 |
| T1D no HLA | Meta Analysis | 1.10 (1.00-1.20) | 0.05 | |
| T2D | UKBB ICD-10 | 1.00 (1.00-1.00) | 0.54 | 0.90 |
| T2D | UKBB GP | 1.01 (0.97-1.06) | 0.59 | 0.22 |
| T2D | FinnGen | 1.01 (0.84-1.22) | 0.87 | 0.73 |
| T2D | Meta Analysis | 1.00 (1.00-1.00) | 0.92 | |
